# The effect of multiple interventions to balance healthcare demand for controlling COVID-19 outbreaks: a modelling study

**DOI:** 10.1101/2020.05.19.20107326

**Authors:** Po Yang, Jun Qi, Shuhao Zhang, Xulong Wang, Gaoshan Bi, Yun Yang, Bin Sheng, Xuxin Mad

## Abstract

**Background:** Recent outbreak of a novel coronavirus disease 2019 (COVID-19) has led a rapid global spread around the world. For controlling COVID-19 outbreaks, many countries have implemented two non-pharmaceutical interventions: suppression like immediate lock-downs in cities at epicentre of outbreak; or mitigation that slows down but not stopping epidemic for reducing peak healthcare demand. Both interventions have apparent pros and cons; the effectiveness of any one intervention in isolation is limited. It is crucial but hard to know how and when to take which level of interventions tailored to the specific situation in each country. We aimed to conduct a feasibility study for robustly accessing the effect of multiple interventions to control the number and distribution of infections, growth of deaths, peaks and lengths of COVID-19 breakouts in the UK and other European countries, accounting for balance of healthcare demand.

**Methods:** We developed a model to attempt to infer the impact of mitigation, suppression and multiple rolling interventions for controlling COVID-19 outbreaks in the UK. Our model assumed that each intervention has equivalent effect on the reproduction number R across countries and over time; where its intensity was presented by average-number contacts with susceptible individuals as infectious individuals; early immediate intensive intervention led to increased health need and social anxiety. We considered two important features: direct link between Exposed and Recovered population, and practical healthcare demand by separation of infections into mild, moderate and critical cases. Our model was fitted and calibrated with date on cases of COVID-19 in Wuhan to estimate how suppression intervention impacted on the number and distribution of infections, growth of deaths over time during January 2020, and April 2020. We combined the calibrated model with data on the cases of COVID-19 in London and non-London regions in the UK during February 2020 and April 2020 to estimate the number and distribution of infections, growth of deaths, and healthcare demand by using multiple interventions. We applied the calibrated model to the prediction of infection and healthcare resource changes in other 6 European countries based on actual measures they have implemented during this period.

**Findings:** We estimated given that 1) By the date (5^th^ March 2020) of the first report death in the UK, around 7499 people would have already been infected with the virus. After taking suppression on 23^rd^ March, the peak of infection in the UK would have occurred between 28^th^ March and 4^th^ April 2020; the peak of death would have occurred between 18^th^ April and 24^th^ April 2020. 2) By 29^th^ April, no significant collapse of health system in the UK have occurred, where there have been sufficient hospital beds for severe and critical cases. But in the Europe, Italy, Spain and France have experienced a 3 weeks period of shortage of hospital beds for severe and critical cases, leading to many deaths outside hospitals. 3) One optimal strategy to control COVID-19 outbreaks in the UK is to take region-level specific intervention. If taking suppression with very high intensity in London from 23^rd^ March 2020 for 100 days, and 3 weeks rolling intervention between very high intensity and high intensity in non-London regions. The total infections and deaths in the UK were limited to 9.3 million and 143 thousand; the peak time of healthcare demand was due to the 96^th^ day (12^th^ May, 2020), where it needs hospital beds for 68.9 thousand severe and critical cases. 4) If taking a simultaneous 3 weeks rolling intervention between very high intensity and high intensity in all regions of the UK, the total infections and deaths increased slightly to 10 million and 154 thousand; the peak time of healthcare occurs at the 97^th^ day (13^th^ May, 2020), where it needs equivalent hospital beds for severe and critical cases of 73.5 thousand. 5) If too early releasing intervention intensity above moderate level and simultaneously implemented them in all regions of the UK, there would be a risk of second wave, where the total infections and deaths in the UK possibly reached to 23.4 million and 897 thousand.

**Interpretation:** Considering social and economic costs in controlling COVID-19 outbreaks, long-term suppression is not economically viable. Our finding suggests that rolling intervention is an optimal strategy to effectively and efficiently control COVID-19 outbreaks in the UK and potential other countries for balancing healthcare demand and morality ratio. As for huge difference of population density and social distancing between different regions in the UK, it is more appropriate to implement regional level specific intervention with varied intensities and maintenance periods. We suggest an intervention strategy to the UK that take a consistent suppression in London for 100 days and 3 weeks rolling intervention in other regions. This strategy would reduce the overall infections and deaths of COVID-19 outbreaks, and balance healthcare demand in the UK.

## Introduction

As of 1^st^ April 2020, the ongoing global epidemic outbreak of coronavirus disease 2019 (COVID-19) has spread to at least 146 countries and territories on 6 continents, resulted in 896 thousands confirmed case and over 45 thousands deaths.^1^ In the UK, COVID-19 infections and deaths reached 29478 and 2352, with a mortality ratio nearly 7.9%.^1^ For effectively controlling COVID19-breaks, most countries have implemented two non-pharmaceutical interventions: suppression strategy like immediate lockdowns in some cities at epicentre of outbreak; or mitigation that slows down but not stopping epidemic for reducing peak healthcare demand.^2.3.4^

However, both above interventions have apparent pros and cons; the effectiveness of any one intervention in isolation is limited.^4.^ Taking an example of controlling the COVID-19 epidemic in Wuhan, suppression strategy with extremely high intensity (the highest state of emergency) were token by China government from 23^nd^ January 2020 for 50 days, resulting prevention of over 700 thousand national infectious case.^5^ However, China’s first quarter gross domestic product is estimated to a year-on-year contraction to 9 percent.^6^ In most scenarios, it is difficult to conduct an optimal intervention that minimises both growing infections and economic loss in ongoing COVID-19 breakouts.

The effectiveness of intervention strategies is accessed by decline of daily reproduction parameter R_t_, that used to measure a transmission potential of a disease. The R_t_ of COVID-19 is widely estimated within a range of value between 2.5 and 3.^7,8.9.10^ Its implementation hinges on two parameters: intervention intensity presented by average-number contacts per person, and intervention duration counted by weeks.^11^ The practical impacts of applying intervention strategies to certain country are varied in light of many factors including population density, human mobility, health resources, culture issues, etc. It is crucial but hard to know how and when to take which level of interventions tailored to the specific situation in each country.

Targeting at this problem, we aimed to conduct a feasibility study that explored a range of epidemiological scenarios by taking different intervention strategies on current information about COVID-19 outbreaks in the UK. We assessed the effectiveness of multiple interventions to control outbreaks using a mathematical transmission model accounting for available and required healthcare resources by distinguishing self-recovered populations, infection with mild and critical cases. By varying the intensity, timing point, period and combinations of multiple interventions, we show how viable it is for the UK to minimise the total number of infections and deaths, delay and reduce peak of healthcare demand. We applied the calibrated model to the prediction of infection and healthcare resource changes in other 6 European countries based on actual measures they have implemented during this period.

## Results

### Effectiveness of suppression

As shown in Fig.1, the model reproduced the observed temporal trend of cases within London, non-London and the UK. We estimated that by the date (5^th^ March 2020) of the first report death in the UK, around 7499 people (0.012% of the UK entire population) would have already been infected with the COVID-19. Before lifting measures to intensive suppression on 23^rd^ Mach 2020 (the 46^th^ day), the UK total infections including exposed and infectious populations would actually reach 349,455, nearly up to 0.52% of the UK population. This figure suggests that there were nearly 23 times more infections in the UK than were reported as confirmed case (6650 on 23^rd^ March). The infections in London nearly occupied about 22% of the overall UK infections. It meant an exponential growth of total infections between 12^th^ March 2020 and April 1^st^, 2020.

**Figure 1:**
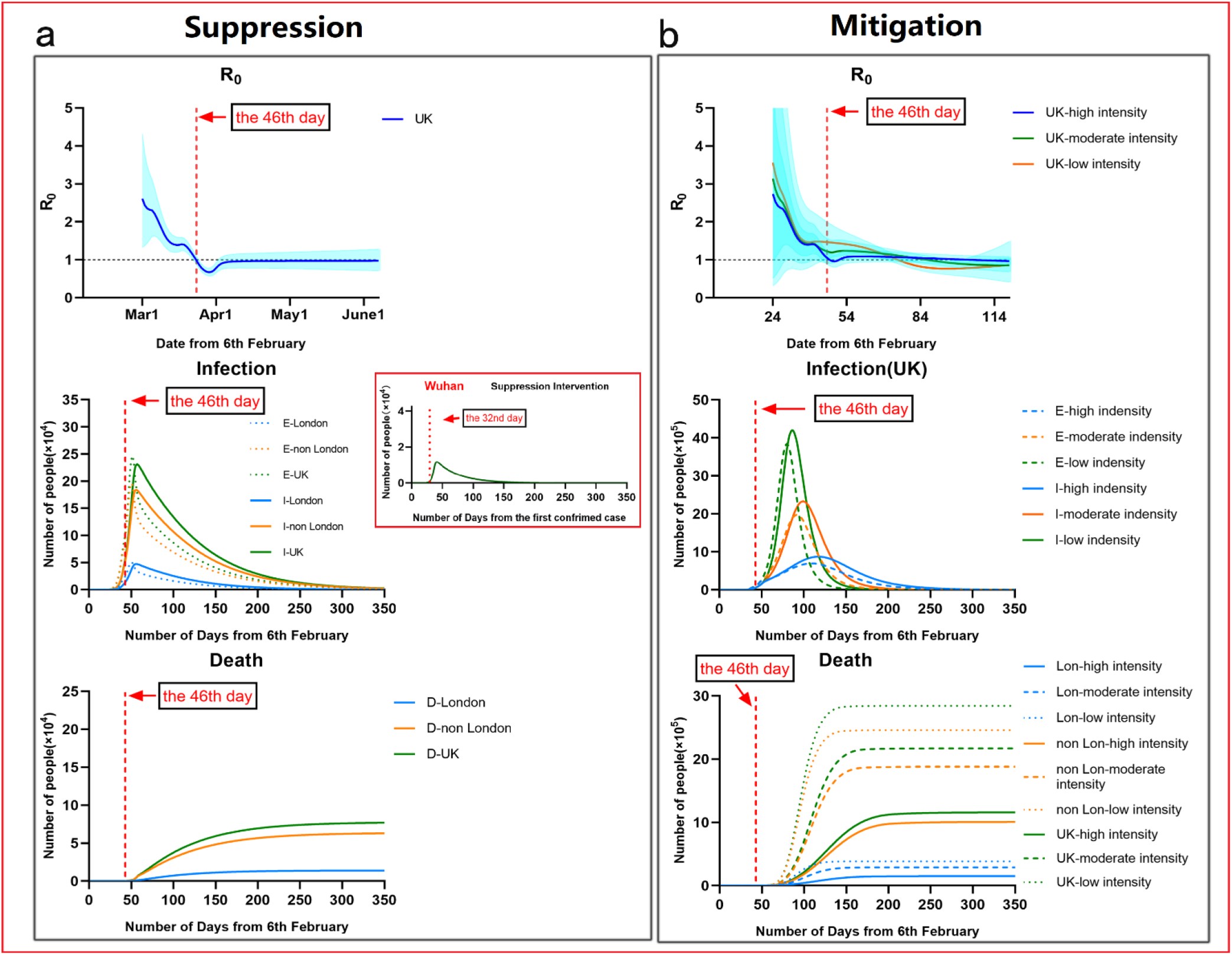
Illustration of controlling COVID-19 outbreaks in London and non-London regions by taking suppression and mitigation with parameters. (a) London population: 9.30 million; non-London population: 57.2 million. (b) Suppression Intervention (M = 3), Mitigation Intervention: Low (M = 10). Moderate (M = 8). High (M =6). (c) Effectiveness of isolation in contact phase (before 12^th^ March 2020): London. 94%, non-London: 88%.

But after taking intensive suppression on 23^rd^ March in the UK, daily exposed and infectious population were greatly reduced. A rapid decline in R has occurred in later March, from 2.61[1.32-4.32] at the 24th day (1st March 2020) to 0.69[0.59-0.79] at the 51st day (28th March 2020). It implied implementing suppression in the UK performed significantly impact on reduction of infections. In Fig.1, we also estimated that the peak of infection in the UK would have occurred between 29^th^ March and 3^rd^ April 2020; the peak of death would have occurred between 18^th^ April and 24^th^ April 2020.

We predicted that if UK could continuously implement insensitive suppression, COVID-19 epidemic would be able to control by 16^th^ May 2020 (the 100^th^ day), and would be nearly ended by 5^th^ July 2020 (the 150^th^ day). In this case, the total deaths by the end on 24^th^ August 2020 in the UK would be about 69511, where London had about 12921 deaths and non-London regions had about 56590 deaths.

In comparing to the prediction at Wuhan using our model, the difference was that the peak of daily infectious population (E = 50200) of London was nearly 1.5 times greater than the one in Wuhan (E = 32880); the peak time (the 50^th^ day) of daily infections in London was 18 days later than the one (the 32^nd^ day) in Wuhan. It was probably because suppression applied in Wuhan (the 32nd day) was 14 days earlier than London (the 46th day). It implied that earlier suppression could reduce infections significantly, but may lead to an earlier peak time of healthcare demand.

### Effectiveness of mitigation

We simulated that mitigation with low, moderate and high intensity (M = 6, 8, 10) were taken in both London and non-London regions in the UK at the 46^th^ day (23^rd^ March 2020), as show in Fig.1. Considering that the UK went to delay phase on the 35^th^ day (12^th^ March 2020), M in the UK was adjusted to 12 from 12^th^ March 2020 to 23^th^ March 2020.

The simulated results showed that mitigation strategies were able to delay the peak of COVID-19 breakouts in the UK but ineffective to reduce total infectious populations. Compared to suppression, mitigation taken in the UK gave a slower decline in R in March, from 2.73[0.97-5.40] on the 24th day (1st March 2020) to 0.98[95% CI 0.88-1.09] on the 110th day (27th May 2020). It implied that during this period, there were still much growth of infections in the UK. But London had lower R than non-London regions.

We estimated that the peak of daily infectious population would increase to 3.6 million (M = 10) to 1.9 million (M = 8) or 0.69 million (M = 6); the peak date of daily infections was about on the 80^th^ (26^th^ April 2020), 92^nd^ (8^th^ May 2020) and 110^th^ day (26^th^ May 2020). Compared to the situation of implementing suppression, the total deaths in the UK would respectively increase to 2.8 million (M = 10) to 2.1 million (M = 8) or 1.1 million (M = 6), where London had about 0.38 million (M = 10) to 0.28 million (M = 8) or 0.15 million (M = 6) and non-London regions had about 2.4 million (M = 10) to 1.8 million (M = 8) or 1 million (M = 6). The periods of COVID-19 epidemic in the UK by taking above mitigations would be extended to over 160, 200 or 300 days.

The result appeared a similar trend as findings,^4^ taking mitigation intervention in the UK enabled reducing impacts of an epidemic by flattening the curve, reducing peak incidence and overall death. While total infectious population may increase over a longer period, the final mortality ratio may be minimised at the end. But as same as taking suppression, mitigation need to remain in place for as much of the epidemic period as possible.

### Health demand and deaths in the UK

From 23^rd^ March 2020, the UK began to implement intensive suppression policy to enforce extreme social distancing. By 29^th^ April 2020, suppression has been implemented for four and half weeks. We used our model to simulate such a period of measure for estimating healthcare demand and deaths in the UK. As shown in Fig.2d, we assumed there were initially 167589 available hospital beds, which was estimated by the number of hospital beds available for every 1000 inhabitants in the UK population.^22^ We assumed at the first day (7^th^ Feb 2020), there are 10% of empty hospital beds (16700) for COVID-19 severe and critical patients. In the first phase of the UK until the 35^th^ day (12^th^ March 2020), government has taken measure to release empty hospital beds from 10% to 12%. Between the 35^th^ Day and the 53th day (3^rd^ April 2020), the action of empty hospital beds in the UK were accelerated, to achieve up to 18.5% of hospital bed availability (31080). After the 53th day, the total number of available hospital beds has sharply risen to 64080. That is because during that time, several Nightingale Hospitals were opened to offer a large number of available beds in the UK.

**Figure 2:**
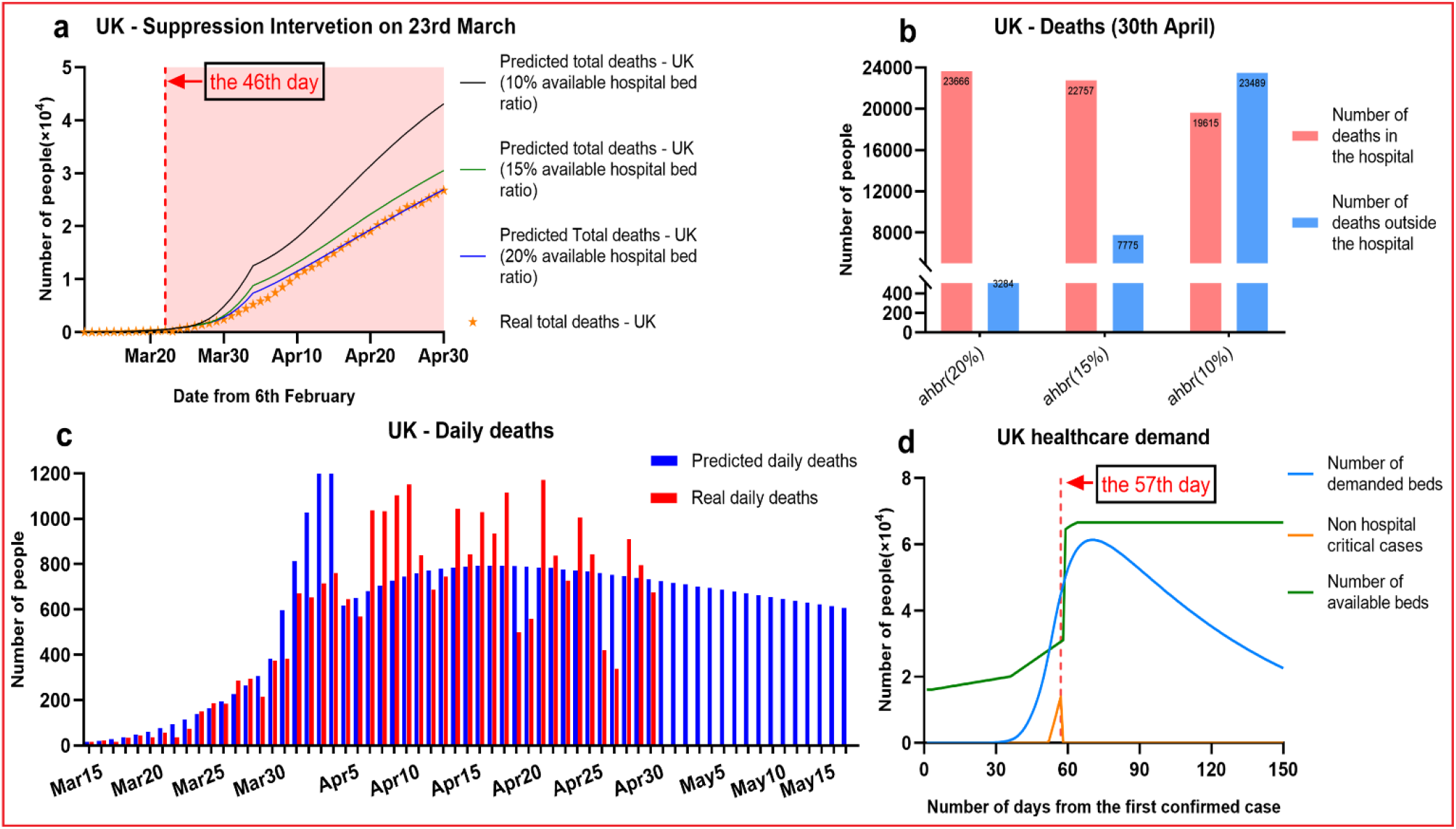
Predicting the impact of Suppression intervention on the UK healthcare demand: a. Forecasting COVID-19 daily deaths in the UK with varied ratio of available hospital bed; b. Forecasting COVID-19 hospital and non-hospital deaths with varied ratio of available hospital bed; c. Demonstration of our predicted daily deaths and real daily deaths by April 30; d. Demonstration of change of the UK healthcare demand over time.

Based on above assumption, as shown in Fig.2a and 2.c, our model accurately predicted the growth of daily death in the UK by 30^th^ April. It appears that the actual number of deaths in the UK on 30^th^ April was 26771. We used the model to predict the total number of deaths in the UK hospital on 30^th^ April is 26950. This figure was on a given assumption that there were around 20% of available hospital beds to supply COVID-19 patients. In this case, all patients can be treated inside the hospital and great reduced the death number outside the hospital. This prediction, shown as an blue line in figure 5(a), is fully fit with current UK real death roll (orange dot line). But in Fig.2b, by 30^th^ April 2020, if the ratio of hospital bed availability reduced to 15%, there would be extra 7,775 non-hospital deaths; if the ratio was lower to 10%, there would be 19615 deaths in the hospital, but more non-hospital deaths 23489. This result revealed that UK government implemented strict admission and discharge criteria to COVID-19 severe and critical patients for protecting NHS. The hospital bed availability continued to be maintained in a good level in preparation of possible second wave.

In the Fig.2d, the blue curve showed our estimated number of demanded hospital beds for COVID-19 severe and critical patients. The blue line demonstrated the change of hospital bed availability over time. The results appeared that at a period between the 50^th^ day (30^th^ March 2020) and 57^th^ day (7^th^ April 2020), there were an amount of non-hospital COVID-19 critical cases, which might lead to increased daily deaths. Expect this period, there were sufficient hospital beds for potential COVID-19 patients. It implied that there were no significant collapse of NHS in the UK.

### Suppression impacts on European countries

We used our model to estimate the impacts of suppression on controlling infections of other 6 EU countries (Italy, Spain, Germany, France, Belgium and Switzerland), as shown in Fig.3. Most suppression in other countries began around 10th-17th March (the 28^th^ – 40^th^ day from first confirmed case). We analyzed data on deaths up to 28th March, giving a 2-3-week window over which to estimate the effect of interventions. For each country, we model the number of infections, the number of deaths, and *R*, the effective reproduction number over time. Specific interventions are assumed to have the same relative impact on *R* in each country when they were introduced there and are informed by mortality data across all countries.

**Figure 3:**
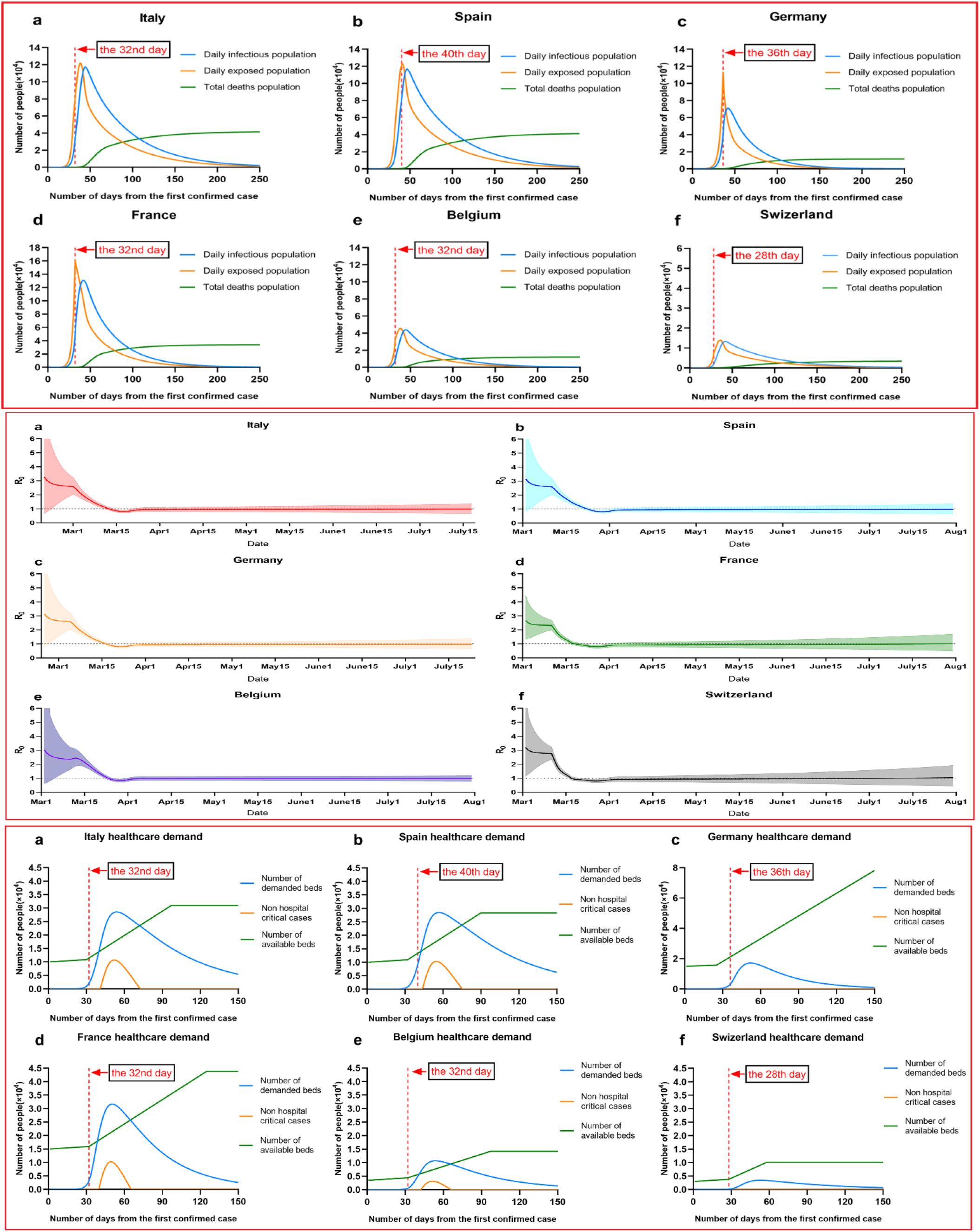
Illustration of forecasting infections, change of reproduction number R, and healthcare demand in 6 European countries (Italy, Spain, Germany, France, Belgium and Switzerland) by implementing suppression intervention.

As shown in Fig.3.a, we made a prediction of the total number of infections and deaths in six European countries. In Italy, our results suggested that, cumulatively, 0.8 [0.684-0.920] million people have been infected as of March 28^th^, giving an attack rate of 9.8% [3.2%-25%] of the population. From 8^th^ February 2020, the total number of infections is about 1.9 million, the total number of deaths is about 41 thousand, the true mortality rate is 2.17%, and the ratio of hospital bed availability for COVID patients is 15%, where it refers to 85% hospital bed occupancy. Spain has seen a similar trend in the number of deaths, and given its smaller population, our model estimates that a higher proportion of the population, 1.15% (0.53 [0.46-0.59] million people) have been infected to date. From 1^st^ February 2020, the total number of infections is 2 million, the number of deaths is 41 thousand, and the true mortality rate is 2.07%. Germany is estimated to have one of the lowest attack rates at 0.51% with 423,193 [348,711-503,599] people infected. From 11^th^ February, the total number of infections is about 0.77 million, and the number of deaths is about 11 thousand. The true mortality rate is 1.5%, and the ratio of hospital bed availability for COVID patients is 20%. In France from 15^th^ February 2020, the total number of infections is about 972,351, and the number of deaths is about 30,532. The true mortality rate is 3.14%, and the ratio of hospital bed availability for COVID patients is 10%. In Belgium from 15^th^ February 2020, the total number of infections is about 991,412 and the number of deaths is about 19,209. The true mortality rate is 1.93%, and the ratio of hospital bed availability for COVID patients is 20%. In Switzerland 19^th^ from February 2020, the total number of infections is 104,109 people, the number of deaths is about 2331 people, the real mortality rate is 2.2%, and the ratio of hospital bed availability for COVID patients is 25%. We estimate that there have been many more infections than are currently reported. The high level of under-ascertainment of infections that we estimate here is likely due to the focus on testing in hospital settings rather than in the community. Despite this, only a small minority of individuals in each country have been infected, with an attack rate on average of 0.92% [0.51%-1.33%] with considerable variation between countries. Our estimates implied that the populations in Europe are not close to herd immunity (−50-75°% if R is 2-4).

Also, Fig.3.a shows total forecasted deaths since the beginning of the epidemic up to and including 30 April under our fitted model. For all above countries, our model fits observed deaths data well (Bayesian goodness of fit tests). We find that, across 6 countries, since the beginning of the epidemic, 104,000 [86,840-122,720] deaths have been averted due to interventions. In Italy and Spain, where the epidemic is advanced, 57,796 [49,184-67,043] and 42,967 [37,166-48982] deaths have been averted, respectively. Even in the UK, which is much earlier in its epidemic, we predict 9,659 [8,065-11,397] deaths have been averted. These numbers give only the deaths averted that would have occurred up to 31 March. If we were to include the deaths of currently infected individuals in both models, which might happen after 30 April, then the deaths averted would be substantially higher.

As shown in Fig.3.b, averaged across all 6 countries, we estimate initial reproduction numbers R of approximately 2.66 [0.80-4.46] -3.27 [0.66-7.91], which is in line with other estimates. Our results, which are driven largely by countries with advanced epidemics and larger numbers of deaths (e.g. Italy, Spain), suggest that these interventions have together had a substantial impact on transmission, as measured by changes in the estimated reproduction number R. Across all countries we find current estimates of R to range from a posterior mean of 0.97 [0.75-1.21] for Italy to a posterior mean of 0.95 [0.72-1.20] for Sweden, with an average of 0.96 across the 6 country posterior means, a 67% reduction compared to the pre-intervention values. Further, with R values dropping substantially, the rate of acquisition of herd immunity will slow down rapidly. This implies that the virus will be able to spread rapidly should interventions be lifted. While the growth in daily deaths has decreased, due to the lag between infections and deaths, continued rises in daily deaths are to be expected for some time. The results suggest that interventions will have a large impact on infections and deaths despite counts of both rising.

In Fig.3,c, we demonstrated change of health demand of six European countries. It showed that in Italy, Spain and France, there were a period of suffering from shortages of available hospital beds, that is, the blue line in the figure (the number of beds required) exceeds the red line (available for COVID-19 patient beds), causing some patients to fail to be hospitalized in time, and the number of patients outside the Yellow Line Hospital has risen. It can be seen that Belgium also has a certain period of shortage of healthcare resources, but compared to the three countries mentioned above, the situation is better with less non-hospital critical cases. The charts of Germany and Switzerland showed that there were no shortage of healthcare resources leading to non-hospital critical cases in both countries. Corresponding to the results shown in Fig.3.a, the shortage of healthcare resources in Italy, Spain and France has caused the death toll to exceed the normal level, making the death rate more than 2%, while the situation in Belgium is lighter. So the real mortality rate is not as high as 1.9%. In Germany and Switzerland, the real mortality rate remains low at around 1.5% owning to enough healthcare resources.

In light of the prediction of our model, it can be seen that the death tolls and death rates in Italy, Spain and France are more than those in Germany, Belgium and Switzerland. The main reason, as presented in Fig.3.c, is the shortage of healthcare resources in Italy, Spain and France leading to high demand for hospital beds than current available beds and will continue for a long time. The other reason is a majority of moderate and severely ill patients are not able to be hospitalized and thus missed the chance to be saved. The highest mortality rate country Italy has higher request on number of hospital beds than their availability during the period of shortage of medical resources.

### Effectiveness of multiple interventions

We simulated two possible situations in London and the UK by implementing rolling interventions as shown in Fig.4. We assumed that all regions in the UK implemented an initial 3 weeks suppression intervention (M=3) from the 46^th^ day (23^rd^ March 2020) to the 67^th^ day (13^rd^ April 2020). Then, two possible rolling interventions were given: 1) to keep suppression in London, and take a 3 weeks rolling intervention between suppression and high intensity mitigation (M = 5) in non-London regions; 2) to take 3 weeks rolling intervention between suppression and high intensity mitigation (M = 5) in all UK.

**Figure 4:**
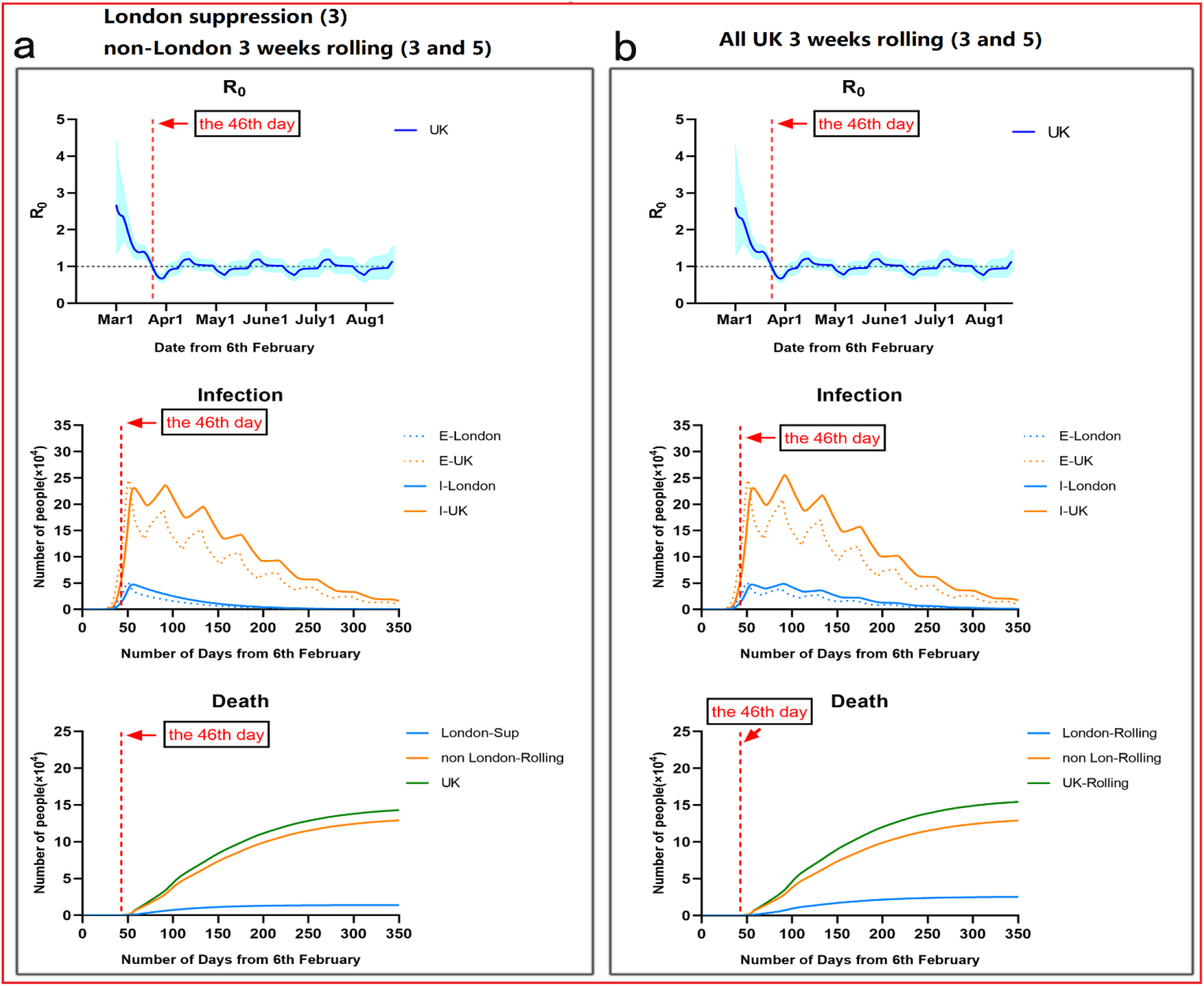
Illustration of controlling COVID-19 outbreaks in London and non-London regions by taking suppression and 3 weeks rolling intervention with parameters. (a) London population: 9.30 million; non-London population: 57.2 million. (b) Suppression Intervention (M = 3), 3 weeks rolling intervention: M = 3-5-3-5, M = 3-4-3-4-3-4. (c) Effectiveness of isolation in contact phase (before 12^th^ March 2020): London. 94%, non-London: 88%.

The simulated results in Fig.4 showed the epidemic appeared a unimodal distribution trend over 350 days, longer than the period of suppression. Similar to suppression in Fig.1, the peak date of infectious population in London or non-London regions remain same at the 50^th^ day. After three weeks, rolling intervention with released intensity in non-London regions led to a fluctuation with 4 or 5 peaks of infections until the end of epidemic. The total deaths and infectious population in the UK were greatly reduced to a range from 143 thousand to 154 thousand. It was about 85% - 100% more than the outcome of taking suppression in all the UK.

Above two rolling interventions taken in the UK gave a similar trend of R as suppression, where there was a fast decline in R in March, from 2.61[1.32-4.32] on the 24^th^ day (1^st^ March 2020) to 0.69[0.59-0.79] on the 51^th^ day (28^th^ March 2020). It implied that 3 weeks rolling intervention (M = 3 or 5) had equivalent effects on controlling transmissions as suppression, but need to be maintained in a longer period of 350 days. From then, R value was oscillated between 1.22 [1.04-1.41] and 0.77[0.63-0.92] with the shrinkage of intervention intensity.

### Optimal rolling intervention

Apart from previous two 3 weeks rolling interventions, we simulated other possible rolling interventions with varied period (2, 3 and 4 weeks) and intervention intensity (M = 4, 5 and 6), as shown in Table. 1: 1) the black part assumed that an initial 3 weeks suppression intervention (M=3) from the 46^th^ day (23^rd^ March 2020) to the 67^th^ day (13^rd^ April 2020) was first implemented in the UK; then after 13^th^ April 2020, other possible rolling interventions were given. 2) the red part assumed that an continues 6 or 9 weeks suppression intervention (M=3) from the 46^th^ day (23^rd^ March 2020) to the 88^th^ day (4^th^ May 2020) or the 109^th^ day (25^th^ May 2020) was first implemented in the UK; then after the 88^th^ or 109^th^ day, other possible rolling interventions were given.

**Table 1:** Performance comparison of rolling interventions in the UK. (FMR: Final morality rate = Total deaths / Total infections. PHSC: Peak value of healthcare demand (Severe and Critical cases), PTSC: Peak time of healthcare demand; PnH: Peak value of non-hospital population, PTnH: Peak time of non-hospital population; TD: Total deaths (UK), TI = Total infections (UK), E: End in 1 year, D: Distribution (Unimodal/Multimodal))

| Multiple interventions<br>(intensity 3-5) | U/M | E | TI | TD | PTSC | PHSC | PTnH | PnH | FMR |
| --- | --- | --- | --- | --- | --- | --- | --- | --- | --- |
| All UK suppression (27W, until 5<sup>th</sup> Oct) M = 3 | U | Y | 5101783 | 76972 | 70 | 61360 | 57 | 13920 | 1.5% |
| All UK 3 weeks rolling ( After 3W suppression,<br>Rolling start from 13<sup>rd</sup> April ) M = 3-4 | U | Y | 6896541 | 102871 | 70 | 61360 | 57 | 13920 | 1.49% |
| All UK 2 weeks rolling ( After 3W suppression,<br>Rolling start from 13<sup>rd</sup> April ) M = 3-5 | U | Y | 10116715 | 160236 | 84 | 73660 | 57 | 13920 | 1.58% |
| All UK 3 weeks rolling ( After 3W suppression,<br>Rolling start from 13<sup>rd</sup> April ) M = 3-5 | U | Y | 10042694 | 154569 | 97 | 73560 | 57 | 13920 | 1.53% |
| All UK 4 weeks rolling ( After 3W suppression,<br>Rolling start from 13<sup>rd</sup> April ) M = 3-5 | U | Y | 9925852 | 151164 | 111 | 72420 | 57 | 13920 | 1.52% |
| All UK 2 weeks rolling ( After 3W suppression,<br>Rolling start from 13<sup>rd</sup> April ) M = 3-6 | M | N | 14159946 | 325904 | 112 | 95650 | 112 | 28990 | 2.30% |
| All UK 3 weeks rolling ( After 3W suppression,<br>Rolling start from 13<sup>rd</sup> April ) M = 3-6 | M | N | 14228064 | 319955 | 139 | 97110 | 139 | 30450 | 2.24% |
| All UK 4 weeks rolling ( After 3W suppression,<br>Rolling start from 13<sup>rd</sup> April ) M = 3-6 | M | N | 14228569 | 310589 | 113 | 101700 | 113 | 35090 | 2.18% |
| All UK 3 weeks rolling ( After 3W suppression,<br>Rolling start from 13<sup>rd</sup> April ) M = 3-8 | M | N | 23351902 | 971622 | 139 | 2287000 | 139 | 162000 | 4.16% |
| London suppression (12W, until 15<sup>th</sup> June) M = 3,<br>other regions 2 weeks rolling ( After 3W<br>suppression, Rolling start from 13<sup>rd</sup> April ) M = 3-5 | U | Y | 9427917 | 147394 | 84 | 70410 | 57 | 13920 | 1.56% |
| London suppression (12W, until 15<sup>th</sup> June) M = 3,<br>other regions 3 weeks rolling ( After 3W<br>suppression, Rolling start from 13<sup>rd</sup> April ) M = 3-5 | U | Y | 9367882 | 143105 | 96 | 68940 | 57 | 13920 | 1.52% |
| London suppression (12W until 15<sup>th</sup> June) M = 3,<br>other regions 4 weeks rolling (After 3W<br>suppression, Rolling start from 13<sup>rd</sup> April ) M = 3-5 | U | Y | 9268946 | 140900 | 110 | 66660 | 57 | 13920 | 1.52% |
| All UK 2 weeks rolling ( After 6W suppression,<br>Rolling start from 4<sup>th</sup> May ) M = 3-5 | M | N | 8694038 | 128310 | 70 | 61360 | 57 | 13920 | 1.47% |
| All UK 3 weeks rolling ( After 6W suppression,<br>Rolling start from 4<sup>th</sup> May ) M = 3-5 | U | Y | 8951775 | 132121 | 70 | 61360 | 57 | 13920 | 1.47% |
| All UK 4 weeks rolling ( After 6W suppression,<br>Rolling start from 4<sup>th</sup> May ) M = 3-5 | U | Y | 9201486 | 135590 | 70 | 61360 | 57 | 13920 | 1.47% |
| All UK 2 weeks rolling ( After 9W suppression,<br>Rolling start from 25<sup>th</sup> May ) M = 3-5 | U | Y | 7761765 | 114503 | 70 | 61360 | 57 | 13920 | 1.47% |
| All UK 3 weeks rolling ( After 9W suppression,<br>Rolling start from 25<sup>th</sup> May ) M = 3-5 | U | Y | 7980859 | 117608 | 70 | 61360 | 57 | 13920 | 1.47% |
| All UK 4 weeks rolling ( After 9W suppression,<br>Rolling start from 25<sup>th</sup> May ) M = 3-5 | U | Y | 8166998 | 120494 | 70 | 61360 | 57 | 13920 | 1.47% |

The results in the first scenario revealed that rolling intervention with middle intensity (M = 6) cannot control the outbreaks in one year, where the distribution of epidemic was a multimodal trend as similar to mitigation outcomes. The overall infections and deaths significantly increased to over 14 million and 268 thousand. While the peak time of healthcare demand for severe critical cases delayed to the 112^nd^ – 139^th^ day, the total deaths of the UK would be double than other rolling interventions with low intensity. Another finding was that given equivalent intensity (M= 3 and 5) of rolling interventions, the longer period (4 weeks) led to slight reduction of the total deaths to 151,164, compared to 154,569 of 3 weeks rolling and 160,236 of 2 weeks rolling in the UK. The peak time of healthcare demand nearly occurred at same: the 84^th^-111^th^ day; with an equivalent peak value. Thus, in balance of total deaths and human mobility restriction, 3 weeks of period might be a feasible choice.

As shown in Table. 1 and Fig.5, we illustrated the results of the second scenarios that the length of initial suppression was extended to 6 or 9 weeks by 4^th^ or 25^th^ May; and then 3 weeks rolling interventions with adjusted intensity (M = 3-5) were implemented. It indicated that when suppression in the UK was extended to 6 weeks by 4^th^ May, later giving 3 or 4 weeks rolling interventions could reduce total infections to 8951775 or 9201486, and the total deaths to 132121 or 135590. Compared with the first scenario of starting 3 weeks rolling from 13^th^ April, the total infections decreased by 1 million and the total deaths decreased by 15,000. When suppression was extended to 9 weeks by 25^th^ May, the total infections and deaths in the UK had further decreased. As shown in Fig.5, we can find that extending the length of suppression can effectively reduce the overall infections and deaths, and strengthen the effects of multiple interventions.

**Figure 5:**
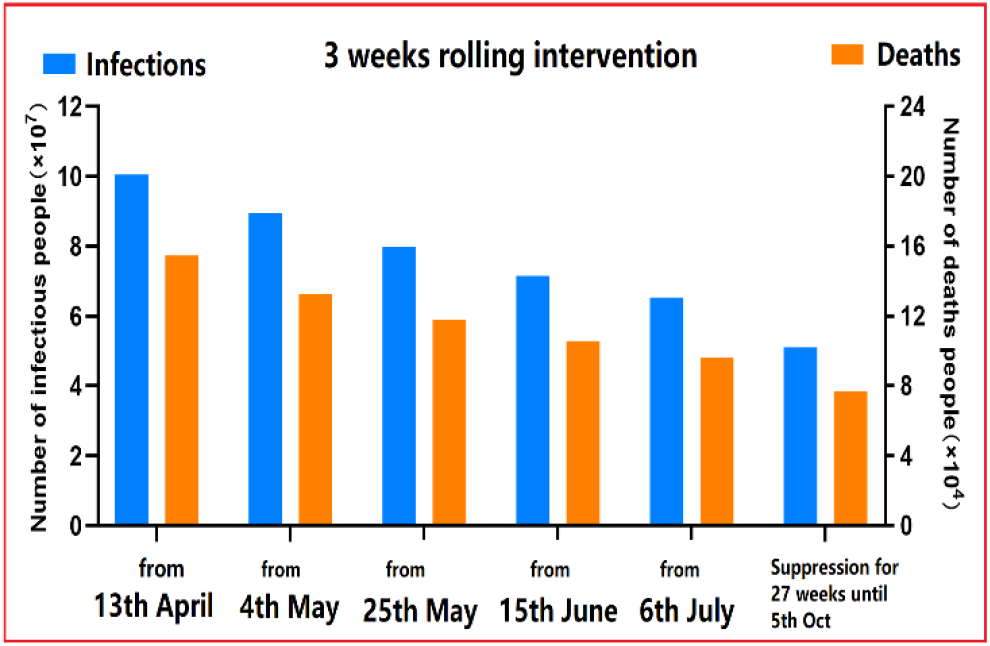
Total infections and deaths in the scenarios of implementing 3 weeks rolling intervention with intensity M = 3 or 5 from different started dates.

**Figure 6:**
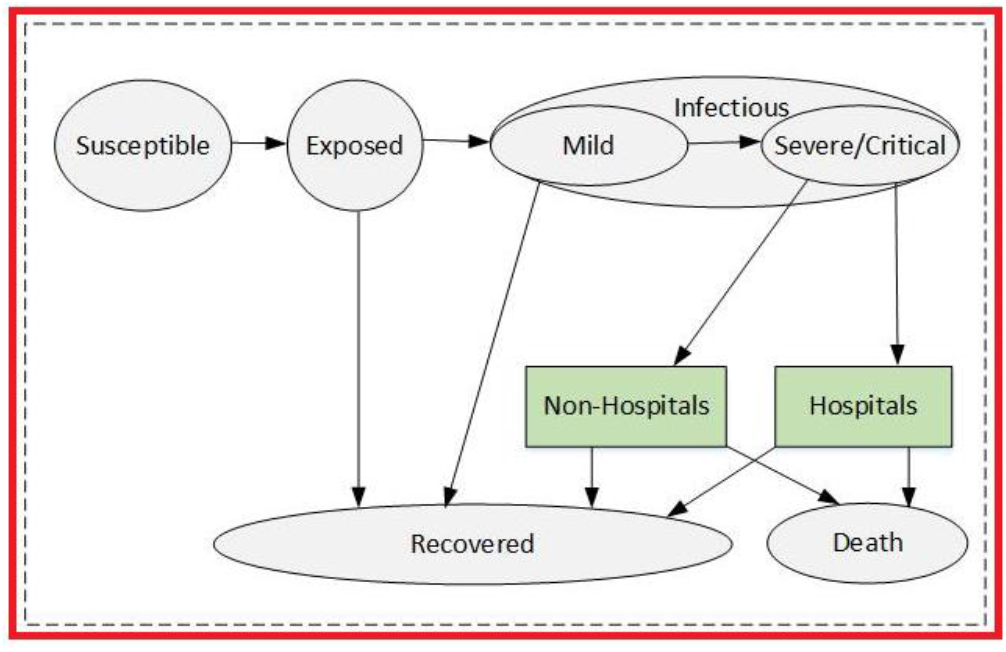
Extended SEMCR model structure. The population is divided into the following six classes: susceptible, exposed (and not yet symptomatic), infectious (symptomatic), mild (mild or moderate symptom), critical (severe symptom), death and recovered (i.e, isolated, recovered, or otherwise non-infectious).

However, the results also indicated that it was possible to control the outbreaks at the 100^th^-150^th^ day that minimized economic loss to the greatest extent. Due to lower population density and less human mobility of non-London regions, 3 weeks rolling intervention was appropriated to non-London regions for balancing the total infections and economic loss, but the length of this strategy was extended to 300 days.

## Discussion

Aiming at a balance of infections, deaths and economic loss, we simulated and evaluated how and when to take which intensity level of interventions was a feasible way to control the COVID-19 outbreak in the Europe. We found rolling intervention between suppression and mitigation with high intensity could be an effective and efficient choice to limit the total deaths but maintain essential mobility for avoiding huge economic lose and society anxiety in a long period. Rolling intervention was more effective in smaller cities. Due to lower population density and less human mobility, realising some intervention intensity would not lead to a second breakout of COVID-19 and benefit maintenance of business activities. Considering difference and diversity of industrial structure of the regions with large population density and small population density in Europe, hybrid intervention was more suitable and effective to control outbreaks. For example, such strategy could complete London outbreak with suppression in 3 months and tolerate a longer recovery period of non-London regions taking 3 weeks rolling intervention. The rapid completion of outbreak in the city like London would strongly benefit to economic recovery. Other regions maintained essential production and business activities to offer sufficient support.

In above scenarios, our model found that the total infections in the UK was limited to 9.3 million; the total deaths in the UK was limited to 143 thousand. Also, the peak time of healthcare demand would occur at the 70^th^ day (16^th^ April 2020), where it needed sufficient hospital beds to accommodate 61.3 thousand severe and critical cases. This scenario echoed that applying suppression at a right time was crucial to delay the peak date of healthcare needs and increase available hospital beds for severe and critical cases. We found that while immediate suppression being taken in Wuhan at 14 days earlier than London reduced 4.7 times infections, it led to nearly 2.34 times of severe and critical cases at non-hospital places (Wuhan: Peak 2789 at the 43^th^ day, London, Peak 1191 at the 57^th^ day). It implied that taking immediate suppression without sufficient hospital beds was risky and led to more deaths in the early breakout.

Our finding revealed that implementing suppression intervention required considering other conditions of this region like culture difference, industrial structure, etc. Success of immediate suppression in Wuhan relied on strict lockdown of human mobility to community level and sufficient resource support from other cities or provinces in China. If there were no sufficiently external support, it would be risky to take highly intensive suppression to entire country due to shortage of healthcare resources and huge impacts on its economics. In Europe, it was hardly to practically implement the same level of intensity as Wuhan. If intensive suppression was relaxed at any time points, the transmission would quickly rebound. This was more like a multi-modal curve when taking multi-intervention strategies in Fig.1. Therefore, we concluded that taking rolling intervention was more suitable to Europe.

Specifically, this control measure could be named as “Besieged and **r**olling **i**nterventions”, that implements hybrid interventions with diverse intensities and different periods of maintenance in region-levels of a country, which measure accounts for each regione intensities and nsity and industrial structure. For many capital cities with high population intensity with closer social distance like Beijing, London, Tokyo, New York, their core businesses are financial service, banking and high technology, which are easily transferred online. Intensive suppression over 2 months plus strict isolation contacts potentially control a second wave of COVID-19 outbreak in city. For other surrounding regions with low population intensity and larger social distancing, 3-4 weeks rolling interventions enable maintaining essential business and production activities, further to provide sufficient support to capital cities. While rolling interventions might last for a longer period, earlier release of capital cities ensures economic recovery of entire country. It is a possible strategy for many other countries to control the first or potential second wave of COVID-19 outbreaks.

Notably, the total infections estimated in our model was measured by Exposed population (asymptomatic), which might be largely greater than other works only estimating Infectious population (symptomatic). We found that a large portion of self-recovered population were asymptomatic or mild symptomatic in the COVID-19 breakouts in Wuhan (occupied about 42%-60% of the total infectious population). These people might think they had been healthy at home because they did not go to hospital for COVID-19 tests. It was one important issue that some SEIR model predicted infectious population in Wuhan that 10 times over than confirmed cases.^12,13^ Early release of intensity might increase a risk of the second breakout.

There are some limitations to our model and analysis. First, our model’s prediction depends on an estimation of intervention intensity that is presented by average-number contacts with susceptible individuals as infectious individuals in a certain region. We assumed that each intervention had equivalent or similar effect on the reproduction number in different regions over time. The practical effectiveness of implementing intervention intensity might be varied with respect to cultures or other issues of certain county. In the UK or similar countries, how to quantify intervention intensity needs an accurate measure of combination of social distancing of the entire population, home isolation of cases and household quarantine of their family members. As for implementing rolling interventions in Europe, the policy needs to be very specific and well-estimated at each day according to the number of confirmed cases, deaths, morality ratio, health resources, etc. Secondly, our model used a variety of plausible biological parameters for COVID-19 based on current evidence as shown in Table.1, but these assumed values might be varied by populations or countries. For instance, we assumed that average period of mild cases to critical cases is 7 days, and average period of elderly people in hospital from severe cases to deaths was 14 days, etc. The change of these variables may impact on our estimation of infections and deaths in the UK. Lastly, our model assumes a condition that there will be a reasonable growth of available hospital source as time goes in the UK after 23^rd^ March 2020. This was actually supported by latest news that Nightingale hospital that enables holding 4000 patients opened at London Excel centre on 4^th^ April 2020.^24^ This assumption is also applicable to several other European countries. As the demand for medical resources continues to expand, the country has begun to expand the available medical resources in its own countries, such as opening temporary tent hospitals, etc.

Our results show that taking rolling intervention is one optimal strategy to effectively and efficiently control COVID-19 outbreaks in the many European countries. This strategy potentially reduces the overall infections and deaths; delays and reduces peak healthcare demand. In future, our model will be extended to investigate how to optimise the timing and strength of intervention to reduce COVID-19 morality and specific healthcare demand.

## Methods

### Mode structure

We implemented a modified SEIR model to account for a dynamic Susceptible [S], Exposed [E] (infected but asymptomatic), Infectious [I] (infected and symptomatic) and Recovered [R] or Dead [D] population s state. For estimating healthcare needs, we categorised infectious group into two sub-cases: Mild [M] and Critical [C]; where Mild cases did not require hospital beds; Critical cases need hospital beds but possibly cannot get it due to shortage of health sources. Conceptually, the modified modal is shown in Figure.5.

The model accounted for delays in symptom onset and reporting by including compartments to reflect transitions between reporting states and disease states. Here, this modal assumed that S is initial susceptible population of certain region; and incorporated an initial intervention of surveillance and isolation of cases in contain phase by a parameter β.^14.15^ If effectiveness of intervention in contain phase was not sufficiently strong, susceptible individuals may contract disease with a given rate when in contact with a portion of exposed population E. After an incubation period α_1_, the exposed individuals became the infectious population I at a ratio 1/α_1_.The incubation period was assumed to be 5.8 days.^8^ Once exposed to infection, infectious population started from Mild cases M to Critical cases C at a ratio *a*, Critical cases led to deaths at a ratio *d*; other infectious population finally recovered. We assumed that COVID-19 can be initially detected in 2 days prior to symptom onset and persist for 7 days in mild cases and 14 days to severe cases.^19^

Notably, two important features in our model differ with other SIR or SEIR models.^12.13^ The first one was that we built two direct relationships between Exposed and Recovered population, Infections with mild symptoms and Recovered population. It was based on an observation of COVID-19 breakouts in Wuhan that a large portion (like 42.5% in Wuhan) of self-recovered population were asymptomatic or mild symptomatic.^14^ They did not go to hospital for official COVID-19 tests but actually were infected. Without considering this issue, the estimation of total infections were greatly underestimated.^13^ In order to measure portion of self-recovery population, we assumed that exposed individuals at home recovered in 3-5 days; mild case at home recovered in 7-10 days.^19^ But if their symptoms get worse, they will be transferred to hospital.

The second feature was to consider shortage of health sources (hospital beds) in the early breakouts of COVID-19 might lead to more deaths, because some severe or critical cases cannot be accommodated in time and led to death at home (non-hospital). For instance, in Wuhan, taking an immediate suppression intervention on 23^rd^ Jan 2020 increased serious society anxiety and led to a higher mortality rate. In order to accurately quantify deaths, our modal considered percentage of elder people in the UK at a ratio occupancy of available NHS hospital beds over time at a ratios H_t_ and their availability for COVID-19 critical cases at a ratio J_t_. We assumed that critical cases at non-hospital places led to death in 4 days; elderly people in critical condition at hospital led to death in 14 days, and non-elderly people in critical condition at hospital led to death in 21 days.^19^

One parameter was defined to measure intervention intensity over time as M_t_. which was presented by average number of contacts per person per day. We assumed that transmission ratio *β* equals to the product of intervention intensity M_t_ and the probability of transmission (b) when exposed (i.e., hospital In Wuhan, intervention intensity was assumed within [3-15], and gave with a relatively accurate estimation of COVID-19 breakouts.^13^

We calibrated its value with respect to the population density and human mobility in London and the UK, and estimated outcomes of COVID-2019 outbreaks by implementing different interventions.

All data and code required to reproduce the analysis is available online at:

https://github.com/TurtleZZH/Comparison-of-Multiple-Interventions-for-Controlling-COVID-19-Outbreaks-in-London-and-the-UK

### Data sources and modal calibration

Considering that COVID-19 breakouts in Wuhan nearly ended by taking suppression intervention, our model was first fitted and calibrated with data on cases of COVID-19 in Wuhan.^13^ In Figure. 1 it showed how suppression (M = 3) impacted on the total number of infections and deaths over time during January 2020 and April 2020. In comparing to other strategies, it demonstrated that the total infections of Wuhan greatly reduced and led an earlier peak time on the 42^nd^ day (2^nd^ Feb 2020. The end time of releasing suppression was due to the 123^th^ day (23^rd^ April 2020). It showed that mitigation (M=6) in Wuhan on the 32^nd^ day may lead to 5-6 times more total infections than suppression, although it would delay the outbreak. If Wuhan took a 2 weeks rolling mitigation and suppression intervention (M = 6 or 3), the total infections might be increased 1.5 times more infections than suppression, although it would delay the outbreak.

Using Wuhan.5 infection our estimation was close to the practical trend of outbreaks in Wuhan, and gave similar results to other works.^13.22^ We tested that transmission rate from I to S is about 0.157; transmission rate from E to S is about 0.787.^13^ The incubation period was assumed to be 6 days.^8^ As for other parameters, we followed the COVID-19 official report from WHO^19^, and gave a medium estimation on average durations related from infectious, to mild or critical case, and death or recovery were shown in Table.2.

**Table 2:**
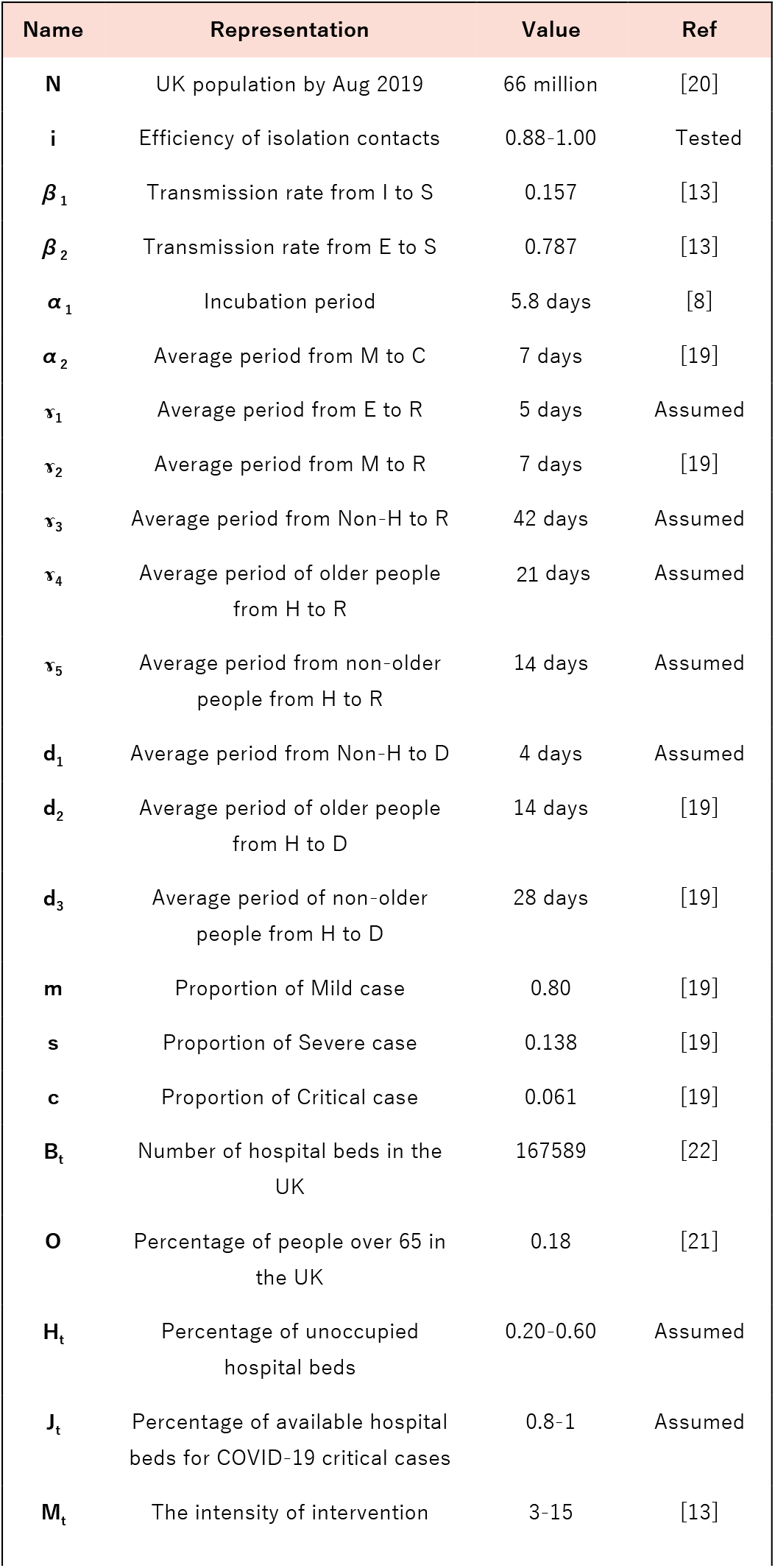
Parameters estimation in our model.

| Name | Representation | Value | Ref |
| --- | --- | --- | --- |
| **N** | UK population by Aug 2019 | 66 million | [20] |
| **i** | Efficiency of isolation contacts | 0.88-1.00 | Tested |
| **$\beta_1$** | Transmission rate from I to S | 0.157 | [13] |
| **$\beta_2$** | Transmission rate from E to S | 0.787 | [13] |
| **$\alpha_1$** | Incubation period | 5.8 days | [8] |
| **$\alpha_2$** | Average period from M to C | 7 days | [19] |
| **$\gamma_1$** | Average period from E to R | 5 days | Assumed |
| **$\gamma_2$** | Average period from M to R | 7 days | [19] |
| **$\gamma_3$** | Average period from Non-H to R | 42 days | Assumed |
| **$\gamma_4$** | Average period of older people from H to R | 21 days | Assumed |
| **$\gamma_5$** | Average period from non-older people from H to R | 14 days | Assumed |
| **$d_1$** | Average period from Non-H to D | 4 days | Assumed |
| **$d_2$** | Average period of older people from H to D | 14 days | [19] |
| **$d_3$** | Average period of non-older people from H to D | 28 days | [19] |
| **m** | Proportion of Mild case | 0.80 | [19] |
| **s** | Proportion of Severe case | 0.138 | [19] |
| **c** | Proportion of Critical case | 0.061 | [19] |
| **$B_t$** | Number of hospital beds in the UK | 167589 | [22] |
| **O** | Percentage of people over 65 in the UK | 0.18 | [21] |
| **$H_t$** | Percentage of unoccupied hospital beds | 0.20-0.60 | Assumed |
| **$J_t$** | Percentage of available hospital beds for COVID-19 critical cases | 0.8-1 | Assumed |
| **$M_t$** | The intensity of intervention | 3-15 | [13] |

Regard as the percentage of elderly people in the UK, it was assumed as 18%.^21^ The total number of NHS hospital beds was given as 167589 with an initial occupied ratio up to 85%.^22^ Considering that UK government began to release NHS hospital beds after COVID-19 breakouts, we assumed the occupied ratio reduced to 80% and would further fall to 40% by 4^th^ April, 2020. Accounting for other serious disease cases requiring NHS hospital beds in the early breakout of COVID-19, we assumed that a ratio of available hospital beds for COVID-19 critical cases was initially at 80%, and gradually raised to 100%.

The intervention intensity was related to the population density and human mobility. We gave an initialization to London and non-London regions: London (M=15, population: 9.3 million), non-London regions (M=14, population: 57.2 million). After taking any kind of interventions, we assumed the change of M would follow a reasonable decline or increase in 3-5 days.

### Procedure

Due to difference of population density between London and other regions in the UK, we observed a fact that the accumulative infections in London was about one third of the total infectious population in the UK.^22^ We separately combined the calibrated model with data on the cases of COVID-19 in London, the UK (non-London) and the UK during February 2020 and March 2020 to estimate the total number of infections and deaths, and also peak time and value of healthcare demand by applying different interventions. In contain stage, we assumed a strategy of isolation contacts were taken in the UK from 6^th^ Feb 2020 to 12^th^ March 2020, the effectiveness of isolation of cases and contacts was assumed as 78% in London and 91% in non-London regions.

The key tuning operation was to adjust intensity level of Mt over time. We assumed that suppression intensity was given to reduce unaltered internal mobility of a region, where: M = 3. Mitigation intensity was given a wide given range [4-12], where high intensity (M = 4 or 5), moderate intensity (M = 6-8), low intensity (M = 912), We evaluated effectiveness of multiple interventions in London and non-London regions, including: suppression, mitigation and rolling intervention. The evaluation metric included 9 indicators as follow: 1: Unimodal or multimodal distribution. 2 If outbreak ends in one year. 3. Total infections. 4. Total deaths. 5. Peak time of healthcare demand. 6. Peak value of healthcare demand for severe and critical cases. 7. Peak time of non-hospital population. 8. Peak value of non-hospital population. 9. Final morality rate (equals to Total deaths over Total infections). The length of intervention was calculated due the date that daily new infections were nearly clear.

Respect to definition of optimal interventions, we first conducted a condition that COVID-19 outbreaks ended as early as possible, and definitely not lasted over 1 year, otherwise it consistently impacted on economic recovery. The second condition was a good balance between total Infections or deaths and intervention intensity. The last one was later peak time and smaller peak value of healthcare demand, where it gave sufficient time to prepare essential health sources.

## Data Availability

All data and code required to reproduce the analysis is available online at:
https://github.com/TurtleZZH/Comparison-of-Multiple-Interventions-for-Controlling-COVID-19-Outbreaks-in-London-and-the-UK

https://github.com/TurtleZZH/Comparison-of-Multiple

## Supplementary Materials

### Transmission model structure

We estimated changes in COVID-19 transmissibility over time via the effective reproduction number (*R_t_*), which represents the mean number of secondary infections that result from a primary case of infection at time t. Values of *R_t_* exceeding 1 indicate that the epidemic will tend to grow, whereas values below 1 indicate that the epidemic will tend to decline. We estimated the time-varying reproduction numbers from serial intervals and incidence of COVID-19 cases over time. The transmission rate β(t) is related to the basic reproduction number by the formula

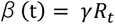

We used particle filter simulation to fit the real (R_t_), the specific steps are:

First step:

a. Generate random initial particles according to the initial state-particle initialization(Our initial R0=3),generally generated using Gaussian random distribution
b. Calculate the initial measurement value from the initial state according to the measurement function equation Second step(Enter the iteration period):
c. Calculate the current state from the initial state according to the state transition matrix or equation
d. Calculate the current observation value from the current state according to the measurement state matrix or equation
e. Generate particles based on the number of particles(We set NN=1000):

1. Generate particles based on the number of particles
2. Calculate the current observation value of the particle according to the measurement function matrix or equation
3. Calculate the likelihood function value to get the particle weight value
f. Normalize the weights of all particles(so that the weight of all particles are between 0-1)
g. Random importance resamples all particles to obtain subscripts with random numbers greater than the limit of particle weights
h. Reassign the current particle sample to the new particle based on the subscript
i. The sate is estimated as the mean of the new particles

We implemented a modified SEIR model to account for a dynamic Susceptible [S], Exposed [E] (infected but asymptomatic), Infectious [I] (infected and symptomatic) and Recovered [R] or Dead [D] population’s state. In order to estimate health demand, we categorized the infectious group into two sub-cases: Mild [M] and Critical [C]; where Mild cases do not need hospital beds; Critical cases need hospital beds but possibly cannot get it due to shortage of health sources.

Following previous assumptions, the implementation of dynamic transmission of our modified SEIR model follows steps as below:

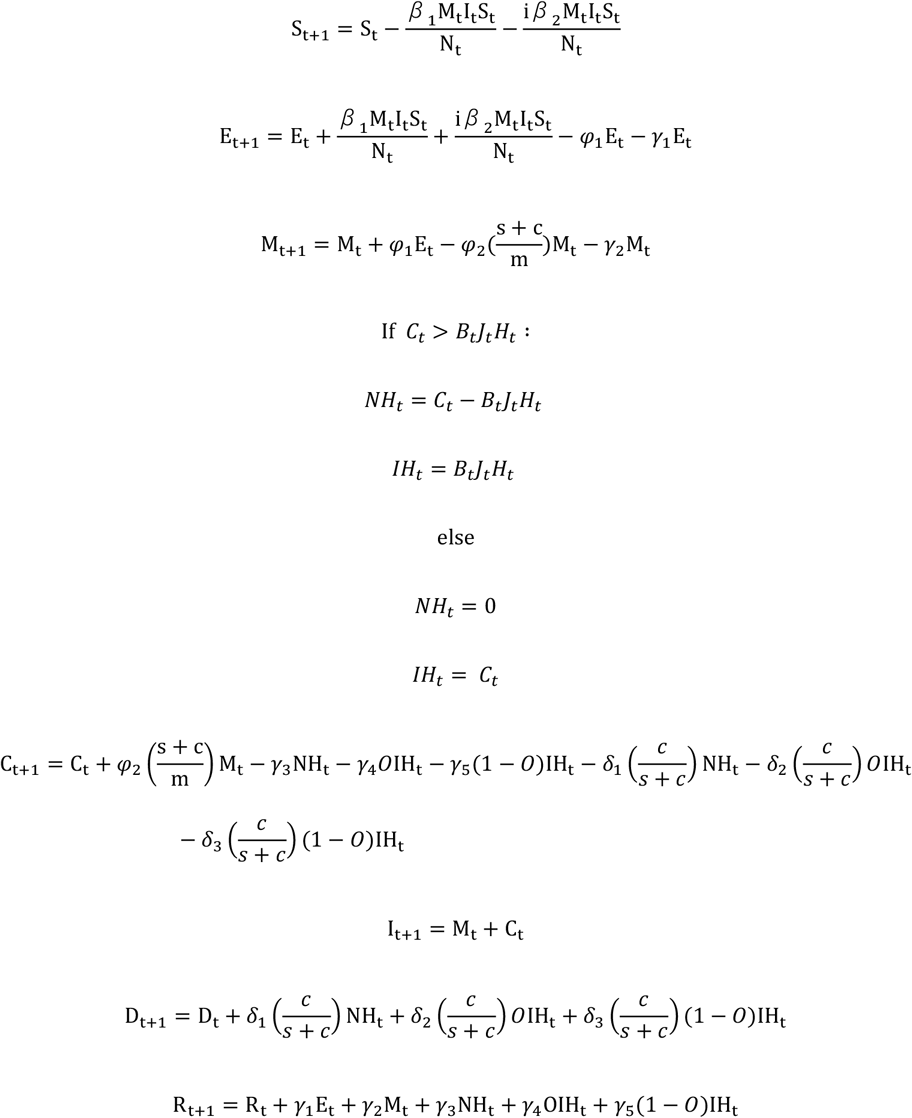

Here S(t) is the number of individuals in UK susceptible at time t, E(t) is the number of people in the UK who have been infected but asymptomatic, I(t) is the number of people in the UK who have been infected and symptomatic, M(t) is the number of people with mild disease, C(t) is the number of people with severe and critical disease, and NH(t) is the number of people with severe and critical disease who have not been hospitalized, IN(t) is the number of people with severe and critical disease who have been hospitalized, R(t) is the number of patients who have been cured, D(t) is the number of patients who have died.

Parameter i is the efficiency of isolation contacts. Parameter m is the proportion of mild case, parameter s is the proportion of severe case, and parameter c is the proportion of critical case. Parameter O is the percentage of people over 65 in the UK.

Parameter *β*_1_ is the transmission rate from I to S, Parameter *β*_2_ is the transmission rate from E to S. Parameter *φ*_1_ is the transmission rate from E to M (1/ α_1_(incubation period)), Parameter *φ*_2_ is the transmission rate from M to C (1/ *α*_2_(average period from M to C)).

Parameter *γ*_1_ is the transmission rate from E to R (1/ ɤ_1_(average period from E to R)), parameter *γ*_2_ is the transmission rate from M to R (1/ ɤ_2_ (average period from M to R)), parameter *γ*_3_ is the transmission rate from NH to R (1/ ɤ_3_ (average period from NH to R)), parameter *γ*_4_ is the transmission rate of older people from IH to R (1/ ɤ_4_(average period of older people from IH to R)), parameter *γ*_5_ is the transmission rate of non-older people from IH to R (1/ ɤ_5_ (average period of non-older people from IH to R)).

Parameter *δ*_1_ is the transmission rate from NH to R (1/ *d*_1_(average period from NH to D)), parameter *δ*_2_ is the transmission rate of older people from IH to R (1/ *d*_2_(average period of older people from IH to D)), parameter *δ*_3_ is the transmission rate of non-older people from IH to R (1/ *d*_3_(average period of non-older people from IH to D)).

Parameter *B_t_* is the number of hospital beds in the UK, parameter *J_t_* is the percentage of available hospital beds for COVID-19 critical cases, *H_t_* is the percentage of unoccupied hospital beds, *M_t_* is the intensity of intervention.

### Data and code availability

All data and code required to reproduce the analysis is available at:

### Table of data on multiple interventions in the UK

Days: Number of days from February 6^th^. Q: The number of total infected population. Z: The number of accumulative infected population. E: The number of daily exposed population. RI: The number of real infected population. D: The number of total dead population. Rate: Mortality rate. PTnH: Peak time of non-hospital population. PnH: Peak value of non-hospital population. Rt: Basic reproduction number)

**Table 1:**
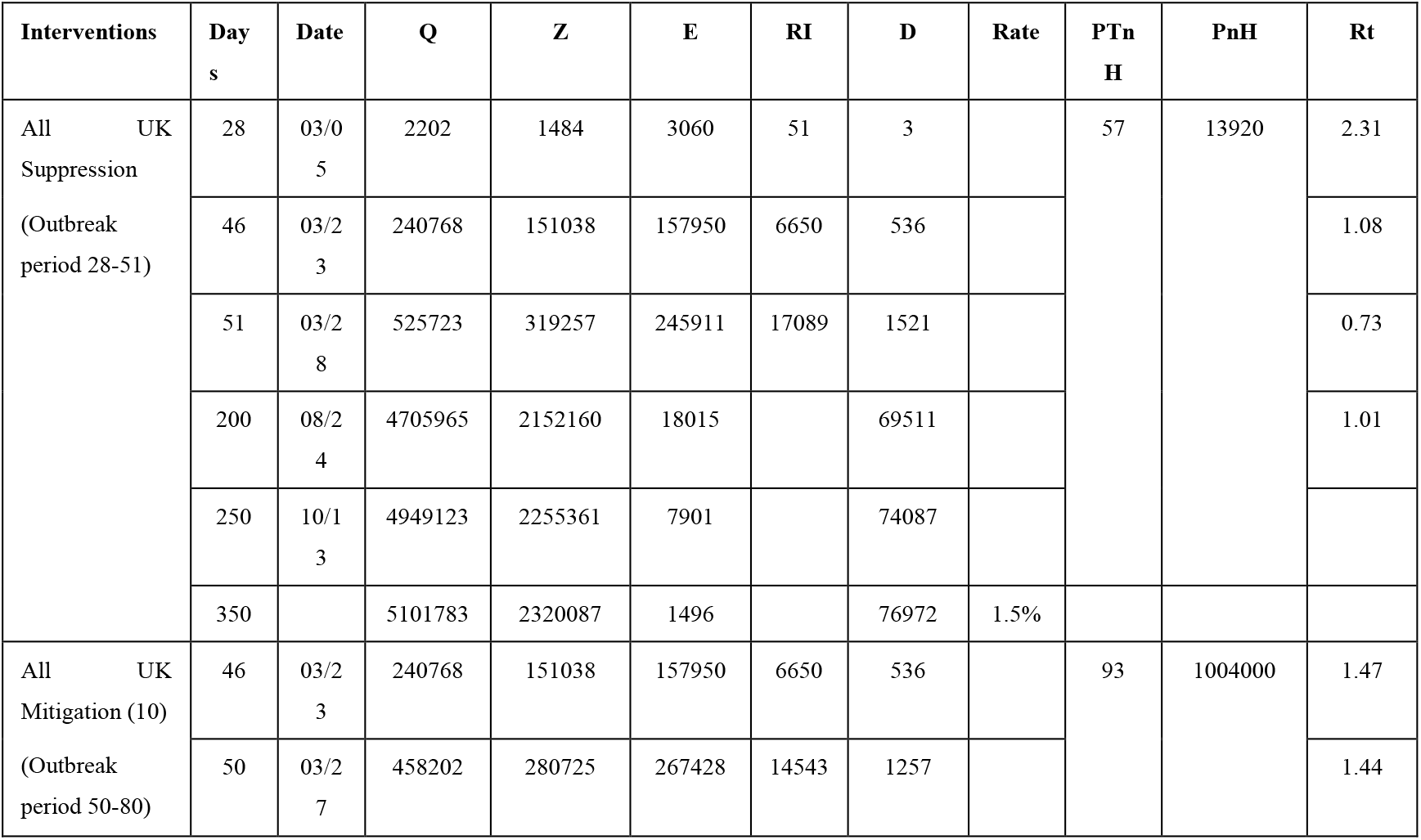
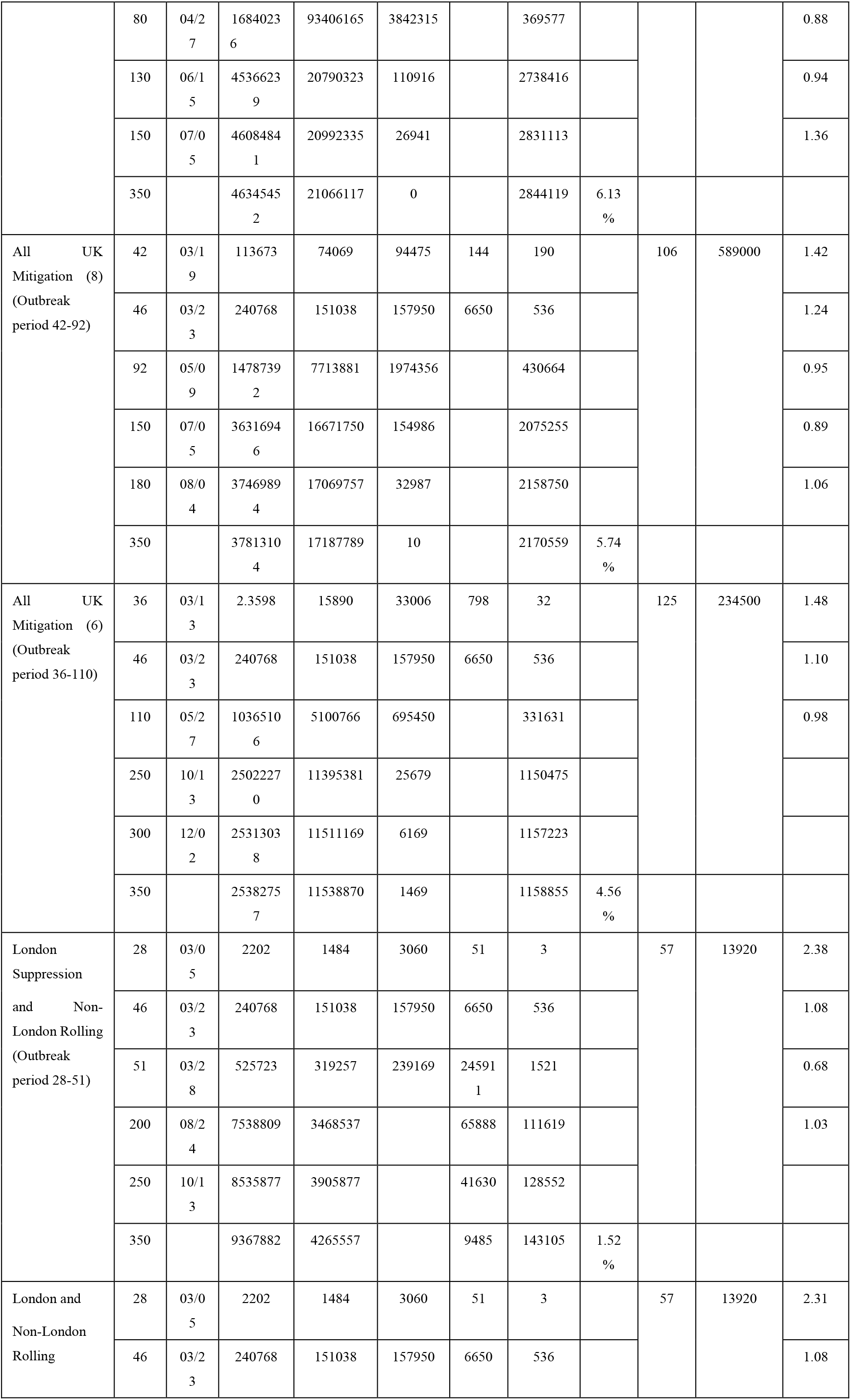
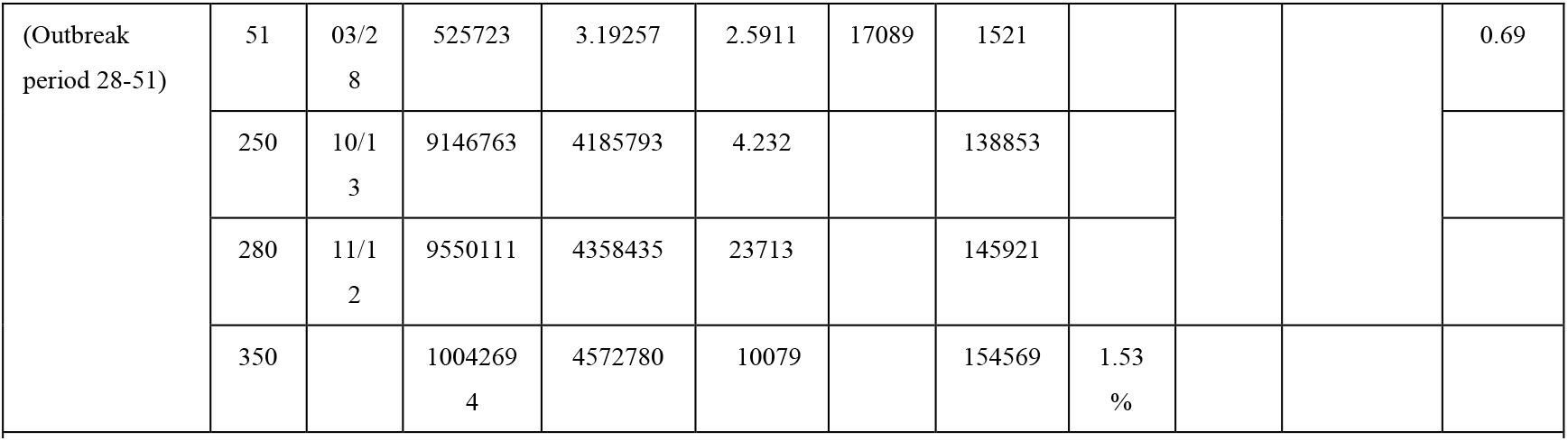
Data on different measures in the UK. (Days: Days from February 6^th^.Q: The number of total infected population. Z: The number of accumulative infected population. E: The number of daily exposed population. RI: The number of real infected population. D: The number of total dead population. Rate: Mortality rate. PTnH: Peak time of non-hospital population. PnH: Peak value of non-hospital population. Rt: Basic reproduction number)

### Table of data on different strong intervention times in the UK

**Table 2:** Data on different suppression intervention times in the UK. (FMR: Final morality rate = Total deaths / Total infections. PHSC: Peak value of healthcare demand (Severe and Critical cases), PTSC: Peak time of healthcare demand; PnH: Peak value of non-hospital population, PTnH: Peak time of non-hospital population; TD: Total deaths (UK), TI = Total infections (UK), E: End in 1 year, D: Distribution (Unimodal/Multimodal))

| Multiple interventions | U/M | E | TI | TD | PTSC | PHSC | PHnT | PnH | FMR |
| --- | --- | --- | --- | --- | --- | --- | --- | --- | --- |
| All UK suppression | U | Y | 5101783 | 76972 | 70 | 61360 | 57 | 13920 | 1.5% |
| All UK 3 weeks rolling (3 and 4) | U | Y | 6896541 | 102871 | 70 | 61360 | 57 | 13920 | 1.49% |
| All UK 2 weeks rolling (3 and 5) | U | Y | 10116715 | 160236 | 84 | 73660 | 57 | 13920 | 1.58% |
| All UK 3 weeks rolling (3 and 5) | U | Y | 10042694 | 154569 | 97 | 73560 | 57 | 13920 | 1.53% |
| All UK 4 weeks rolling (3 and 5) | U | Y | 9925852 | 151164 | 111 | 72420 | 57 | 13920 | 1.52% |
| All UK 2 weeks rolling (3 and 6) | M | N | 14159946 | 325904 | 112 | 95650 | 112 | 28990 | 2.30% |
| All UK 3 weeks rolling (3 and 6) | M | N | 14228064 | 319955 | 139 | 97110 | 139 | 30450 | 2.24% |
| All UK 4 weeks rolling (3 and 6) | M | N | 14228569 | 310589 | 113 | 101700 | 113 | 35090 | 2.18% |
| All UK 3 weeks rolling (3 and 8) | M | N | 23351902 | 971622 | 139 | 2287000 | 139 | 162000 | 4.16% |
| London suppression (3), other regions 2 weeks rolling (3 and 5) | U | Y | 9427917 | 147394 | 84 | 70410 | 57 | 13920 | 1.56% |
| London suppression (3), other regions 3 weeks rolling (3 and 5) | U | Y | 9367882 | 143105 | 96 | 68940 | 57 | 13920 | 1.52% |
| London suppression (3), other regions 4 weeks rolling (3 and 5) | U | Y | 9268946 | 140900 | 110 | 66660 | 57 | 13920 | 1.52% |
| London All suppression -- nonLondon 3-5 3 weeks rolling | U | Y | 8441584 | 124757 | 70 | 61360 | 57 | 13920 | 1.47% |
| Six weeks suppression,then high intensity mitigation 3-5 | M | N | 15622092 | 302186 | 184 | 82650 | 187 | 16380 | 1.93% |
| Six weeks suppression,then high intensity mitigation 3-6 | M | N | 23685805 | 878121 | 180 | 173500 | 180 | 106800 | 3.70% |
| Six weeks suppression,then moderate intensity mitigation 3-7 | M | N | 30647196 | 1451931 | 167 | 314100 | 167 | 247500 | 4.73% |
| Six weeks suppression,then moderate intensity mitigation 3-8 | M | N | 36440443 | 1929340 | 155 | 488600 | 155 | 422000 | 5.29% |
| Six weeks suppression,then 3 week rolling 3-5 | U | Y | 8951775 | 132121 | 70 | 61360 | 57 | 13920 | 1.47% |
| Six weeks suppression,then 3 week rolling 3-6 | M | N | 13092598 | 210152 | 161 | 77220 | 57 | 13920 | 1.60% |
| Six weeks suppression,then 3 week rolling 3-7 | M | N | 17696613 | 467888 | 161 | 121600 | 161 | 54950 | 2.64% |
| Six weeks suppression,then 3 week rolling 3-8 | M | N | 22339206 | 816746 | 161 | 129800 | 161 | 126100 | 3.65% |
| Six weeks suppression,then 2 week rolling 3-5 | M | N | 8694038 | 128310 | 70 | 61360 | 57 | 13920 | 1.47% |
| Six weeks suppression,then 2 week rolling 3-6 | M | N | 12632168 | 187928 | 167 | 68290 | 57 | 13920 | 1.48% |
| Six weeks suppression,then 2 week rolling 3-7 | M | N | 17078493 | 409394 | 168 | 104500 | 168 | 37860 | 2.39% |
| Six weeks suppression,then 2 week rolling 3-8 | M | N | 21555482 | 736153 | 168 | 160100 | 168 | 93430 | 3.41% |
| Six weeks suppression,then 4 week rolling 3-5 | U | Y | 9201486 | 135590 | 70 | 61360 | 57 | 13920 | 1.47% |
| Six weeks suppression,then 4 week rolling 3-6 | M | N | 13468065 | 238766 | 128 | 84680 | 128 | 18020 | 1.77% |
| Six weeks suppression,then 4 week rolling 3-7 | M | N | 18204675 | 521281 | 181 | 132000 | 181 | 65330 | 2.86% |
| Six weeks suppression,then 4 week rolling 3-8 | M | N | 22937138 | 882755 | 181 | 1960000 | 181 | 129400 | 3.84% |
| Nine weeks suppression,then 2 week rolling 3-5 | U | Y | 7761765 | 114503 | 70 | 61360 | 57 | 13920 | 1.47% |
| Nine weeks suppression,then 3 week rolling 3-5 | U | Y | 7980859 | 117608 | 70 | 61360 | 57 | 13920 | 1.47% |
| Nine weeks suppression,then 4 week rolling 3-5 | U | Y | 8166998 | 120494 | 70 | 61360 | 57 | 13920 | 1.47% |
| Eight weeks suppression,then 2 week rolling 3-5 | U | Y | 8057850 | 118845 | 70 | 61360 | 57 | 13920 | 1.47% |
| Eight weeks suppression,then 3 week rolling 3-5 | U | Y | 8289685 | 122218 | 70 | 61360 | 57 | 13920 | 1.47% |
| Eight weeks suppression,then 4 week rolling 3-5 | U | Y | 8494280 | 125231 | 70 | 61360 | 57 | 13920 | 1.47% |

## Notes

### Competing Interest Statement

The authors have declared no competing interest.

